# Multiethnic genome-wide association study of differentiated thyroid cancer in the EPITHYR consortium

**DOI:** 10.1101/2020.07.19.20153841

**Authors:** Thérèse Truong, Fabienne Lesueur, Pierre-Emmanuel Sugier, Julie Guibon, Constance Xhaard, Mojgan Karimi, Om Kulkarni, Elise Lucotte, Delphine Bacq-Daian, Anne Boland-Auge, Claire Mulot, Pierre Laurent-Puig, Claire Schvartz, Anne-Valérie Guizard, Yan Ren, Elisabeth Adjadj, Frédérique Rachédi, Francoise Borson-Chazot, Rosa Maria Ortiz, Juan J. Lence-Anta, Celia M. Pereda, Daniel F. Comiskey, Huiling He, Sandya Liyanarachchi, Albert de la Chapelle, Rosella Elisei, Federica Gemignani, Hauke Thomsen, Asta Forsti, Anthony F. Herzig, Anne-Louise Leutenegger, Carole Rubino, Evgenia Ostroumova, Ausrele Kesminiene, Marie-Christine Boutron-Ruault, Jean-François Deleuze, Pascal Guénel, Florent de Vathaire

## Abstract

Incidence of differentiated thyroid carcinoma (DTC) varies considerably between ethnic groups, with particularly high incidence rates in Pacific Islanders. Here, we conducted a genome-wide association study (GWAS) involving 1,554 cases/1,973 controls of European ancestry and 301 cases/348 controls of Oceanian ancestry from the EPITHYR consortium. Our results confirmed the association with the known DTC susceptibility loci at 2q35, 8p12, 9q22.33 and 14q13.3 in the European ancestry population and suggested two novel signals at 1p31.3 (rs334729) and 16q23.2 (rs16950982), which were associated with TSH levels in previous GWAS. We additionally replicated an association with 5p15.33 reported previously in Chinese and European populations. Except at 1p31.3, all associations were in the same direction in the population of Oceanian ancestry. The frequencies of risk alleles at 2q35, 5p15.33 and 16q23.2 were significantly higher in Oceanians than in Europeans and may explain part of the highest DTC incidence observed in Oceanians.

## INTRODUCTION

The incidence of thyroid cancer was estimated to be 567,233 new cases worldwide in 2018, representing 3.1% of all cancers (1). Papillary thyroid carcinoma (PTC) and follicular thyroid carcinoma (FTC) are the most frequent types of differentiated thyroid carcinomas (DTC) accounting for about 90% of all thyroid cancers. The incidence of DTC is characterized by 3-4 times higher rates in women than in men and wide geographic and ethnic variations (1,2). The highest incidence rates in 2018 were estimated in North America, Pacific Islands and Eastern Asia (1). In some multi-ethnic populations such as in New Caledonia or New Zealand, higher rates were reported in Pacific Islanders than in other ethnic groups (3–5). The causes underlying these geographic and ethnic variations are still unknown and a role for environmental and genetic factors is suspected. During the last decades, the incidence of DTC has increased in most high-resource countries (6).

Part of the increase in incidence has been attributed to changes in medical screening practices that enhance the detection of micro-PTC (≤1 cm in diameter) that are of minimal clinical relevance. It has also been suggested that changes in environmental and lifestyle risk factors may contribute to this increase (7). Apart from exposure to ionizing radiation during childhood, a well-established risk factor for DTC (8), increased DTC risk has also been consistently associated with obesity (9–12). Other risk factors such as deficient or excess iodine intake (13,14) or some reproductive and menstrual factors (15) are suspected to play a role in the etiology of DTC. By contrast, several studies have reported an inverse association between DTC risk and cigarette smoking (16), alcohol intake (17) or oral contraceptive pills (18–20).

DTC is also one of the cancers with the highest familial risk (21), suggesting a major role of genetic risk factors. However, only a few susceptibility loci have been identified thus far by genome-wide association studies (GWAS) (22–29), with limited sample size. All previous GWAS, except one, were conducted in European descent populations and highlighted four susceptibility loci at 2q35, 9q22.33, 8p12 and 14q13.3 that were subsequently widely replicated in diverse populations (30–34). One of these GWAS was conducted in Belarus and focused on radiation-related PTC by including individuals aged under 18 years old at the time of the Chernobyl accident. This study highlighted no other signal than the one at locus 9q22.33. One GWAS was conducted in a population of Asian ancestry in Korea and in addition to the same four previously reported loci identified in populations of European ancestry, reported six other genomic regions at 1p13.3, 1q42.2, 3p14.2, 4q21.1, 12q14.3 and 19q13.2 (29). More recently, a meta-analysis of five GWAS that included 3,001 cases and 287,550 controls of European ancestry (28) reported five new susceptibility loci at 1q42.2, 3q26.2, 5q22.1, 10q24.33 and 15q22.33. However, to our knowledge findings from this meta-analysis have not been replicated in an independent sample.

In order to assess the contribution of known DTC susceptibility loci and to identify new ones in individuals of European ancestry and in individuals of Oceanian ancestry from Pacific Islands, we conducted a multiethnic GWAS in unexplored case-control studies from the EPITHYR consortium.

## RESULTS

### Genome-wide analysis

A total of 1,554 cases and 1,973 controls of European ancestry and 301 case and 348 control individuals of Oceanian ancestry from seven case-control studies were included in the analyses (**Supplementary Table 1**). The study sample included more than 80% women and most of the participants were younger than 50 years old in both ethnic groups. European cases mostly developed PTC, with a higher proportion of tumors larger than 10 mm, while papillary microcarcinomas were more frequent in Oceanian cases (**Table 1**). We performed a GWAS separately in each ethnic group and identified significant variants (i.e. reaching the genome-wide significance P-value threshold of 5×10^−8^) at loci 1p31.3, 2q35, 8p12, 9q22.33 and 14q13.3 in Europeans, and none in Oceanians (**Figure 1**). There was no indication of genomic inflation in both GWAS (λ = 1.02 in Europeans and λ = 1.04 in Oceanians, **Supplementary Figure 1**) suggesting no confounding by cryptic population structure. In the pooled analysis combining both populations (inflation factor λ = 1.08), no additional loci were highlighted and the association at locus 1p31.3 became non-significant (**Supplementary Figure 2**). We also conducted an analysis restricted to PTC cases in Europeans and observed a strengthened association at locus 8p12, but no additional signal was evidenced (**Supplementary Figure 3**).

**Table 1.** Demographic characteristics of the study populations by ethnic group.

|  |  | European (N=3527) |  |  |  | Oceanian (N=649) |  |  |  |
| --- | --- | --- | --- | --- | --- | --- | --- | --- | --- |
|  |  | *Cases* |  | *Controls* |  | *Cases* |  | *Controls* |  |
|  |  | N=1554 | % | N=1973 | % | N=301 | % | N=348 | % |
| **Study** | **Cathy** | 451 | 28.9 | 534 | 27.1 | 1 | 0.3 | 0 |  |
|  | **Young-Thyr** | 637 | 41.0 | 661 | 33.5 | 3 | 1.0 | 0 |  |
|  | **E3N** | 277 | 17.8 | 287 | 14.5 | 0 |  | 0 |  |
|  | **Chernobyl** | 66 | 4.2 | 304 | 15.4 | 0 |  | 0 |  |
|  | **Cuba** | 102 | 6.6 | 113 | 5.7 | 0 |  | 0 |  |
|  | **New Caledonia** | 21 | 1.3 | 70 | 3.5 | 183 | 60.8 | 161 | 46.3 |
|  | **French Polynesia** | 0 | 0.0 | 4 | 0.2 | 114 | 37.9 | 181 | 52.0 |
| **Age**<br>**(years)** | **<25** | 272 | 17.5 | 513 | 26.0 | 0 |  | 0 |  |
|  | **[25-50[** | 763 | 49.1 | 887 | 45.2 | 202 | 67.1 | 255 | 73.3 |
|  | **≥50** | 519 | 33.4 | 573 | 29.0 | 99 | 32.9 | 93 | 26.7 |
| **Sex** | **Women** | 1279 | 82.3 | 1519 | 77.0 | 278 | 92.4 | 318 | 91.4 |
|  | **Men** | 275 | 17.7 | 454 | 23.0 | 23 | 7.6 | 30 | 8.6 |
| **Histology** | **Follicular** | 137 | 8.8 |  |  | 46 | 16.4 |  |  |
|  | **Papillary** | 1417 | 91.2 |  |  | 253 | 83.9 |  |  |
|  | **< 10 mm** | 506 | 35.7 |  |  | 136 | 53.6 |  |  |
|  | **≥ 10 mm** | 904 | 63.8 |  |  | 107 | 42.1 |  |  |

**Figure 1.**
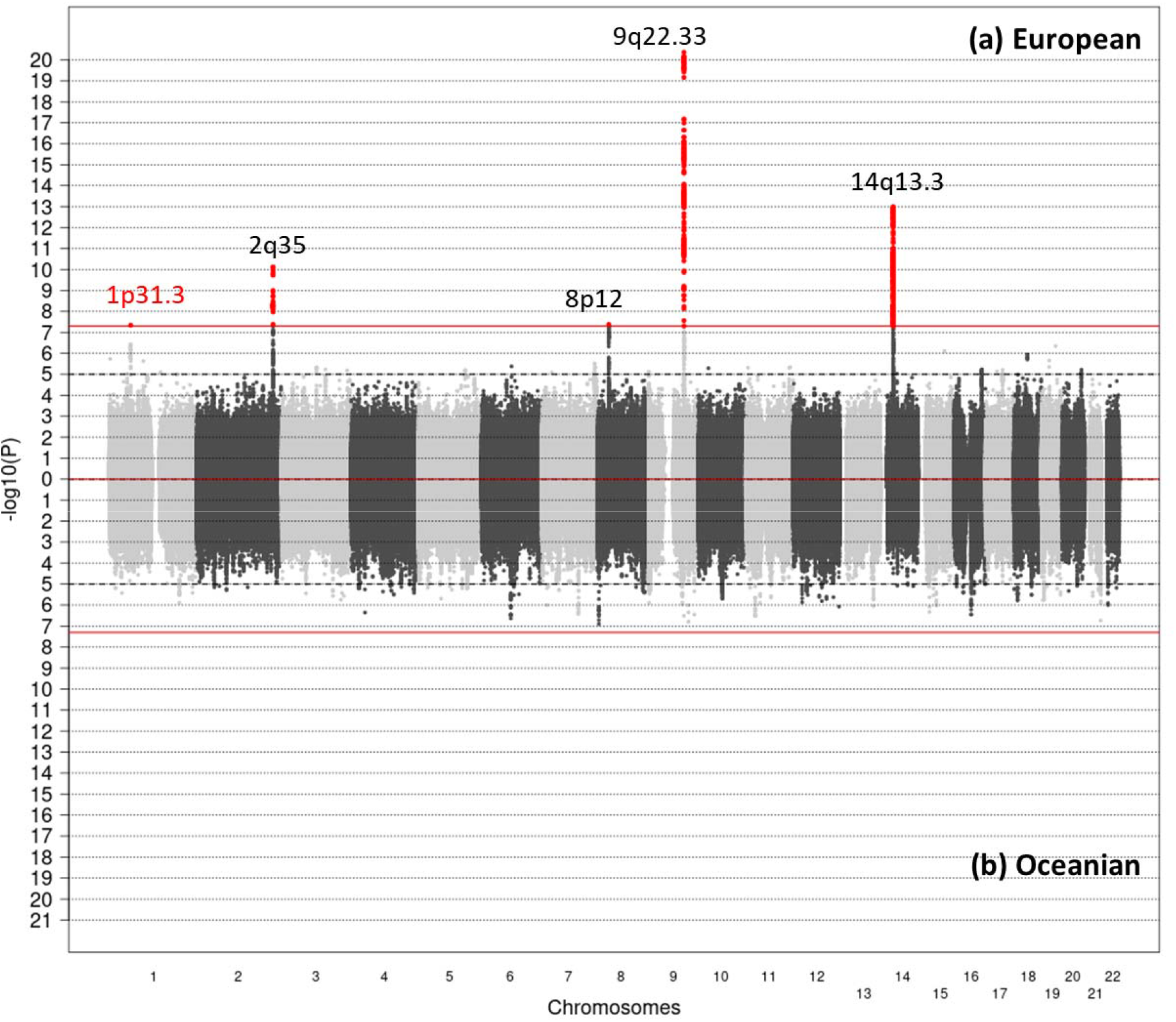
Miami plot of the DTC GWAS results stratified by ethnic group. **(a)** European: 9,673,063 SNPs from 1,554 cases and 1,973 controls. **(b)** Oceanian: 7,179,638 SNPs from 301 cases and 348 controls. The x axis represents chromosomal location and the y axis represents –log_10_ (P-value) from a logistic mixed model (score test). Horizontal dashed lines indicate a genome-wide suggestiveness threshold at p=10^−5^ while red lines indicate genome-wide significance threshold at P=5×10^−8^. Red points denote SNPs that were significantly associated with DTC. Previously published loci are indicated in black and the new locus identified in this analysis is indicated in red.

For each significant locus, we report the association with the SNP with the smallest P-value in the genomic region (lead SNP) according to the European GWAS in **Table 2**. Except at 1p31.3, the associations with these SNPs were in the same direction in Oceanians.

**Table 2.** Association with the lead SNP at each significant locus by ethnic group.

| locus | Lead SNP | Position<br>(hg19) | OA | EA | European |  |  |  | Oceanian |  |  |  |
| --- | --- | --- | --- | --- | --- | --- | --- | --- | --- | --- | --- | --- |
|  |  |  |  |  | EAF | OR | (95%CI) | P | EAF | OR | (95%CI) | P |
| **1p31.3** | **rs334729<sup>(a)</sup>** | 61 608 473 | C | G | 0.95 | **0.53** | (0.41, 0.66) | $8.7 \times 10^{-8}$ <sup>(b)</sup> | 0.86 | **1.02** | (0.72, 1.44) | 0.92 |
| **2q35** | **rs3821098** | 218 292 141 | T | C | 0.72 | **0.70** | (0.62, 0.78) | $1.0 \times 10^{-10}$ | 0.10 | **0.57** | (0.41, 0.81) | $1.4 \times 10^{-3}$ |
| **8p12** | **rs142450470<sup>(a)</sup>** | 32 365 296 | CT | C | 0.35 | **1.33** | (1.19, 1.47) | $1.5 \times 10^{-7}$ <sup>(c)</sup> | 0.22 | **1.18** | (1.11, 1.26) | 0.23 |
| **9q22.33** | **rs1588635** | 100 537 802 | A | C | 0.61 | **0.61** | (0.55, 0.68) | $2.1 \times 10^{-21}$ | 0.72 | **0.75** | (0.58, 0.97) | 0.03 |
| **14q13.3** | **rs368187** | 36 532 576 | C | G | 0.63 | **1.47** | (1.32, 1.63) | $3.8 \times 10^{-13}$ | 0.31 | **1.51** | (1.16, 1.96) | $2.3 \times 10^{-3}$ |
| | **rs116909374** | 36 738 361 | T | C | 0.94 | **0.43** | (0.33, 0.55) | $1.6 \times 10^{-10}$ | 0.99 | **0.24** | (0.05, 1.17) | 0.08 |

Lead SNP: SNP with the smallest P-value in the European GWAS; OA: other allele; EA: effect allele; EAF: effect allele frequency in controls; OR: odds ratio derived from the log-additive logistic mixed model; 95%CI: 95% confidence interval. P: P-value given by the Wald test.
(a) imputed SNP, all imputed SNPs have imputation $r^2 > 0.90$ in both ethnic groups. (b): The P-value given by the score test was $4.5 \times 10^{-8}$ . (c): The P-value given by the score test was $4.2 \times 10^{-8}$ .

### Known loci

DTC susceptibility loci at 2q35, 8p12, 9q22.33 and 14q13.3 have been highlighted in previous GWAS conducted in Europeans. Lead SNPs at 2q35, 9q22.33 and 14q13.3 in the present study were the same or highly correlated with those reported in the published meta-analysis of GWAS on DTC (28) (**Supplementary Table 2**). At 8p12, the SNP rs142450470 in the present study (OR=1.33, 5×10^−7^ for DTC, OR =1.36, P=1.8×10^−8^ for PTC in Europeans) was moderately correlated with rs2439302 highlighted by previous GWAS (22,24) (r^2^ =0.45, OR=1.19, P=4.8×10^−4^ in our study) (**Supplementary Table 2**). The conditional analysis of rs142450470 on rs2439302 gave an OR=1.32 with P =1.3×10^−4^, whereas the conditional analysis of rs2439302 on rs142450470 provided an OR=1.01, P=0.97, suggesting a single signal at this locus.

Stratified analyses by histology showed that the association with rs1588635 at 9q22.33 was stronger for PTC, in particular of size >10 mm, while the association with rs142450470 at 8p12 was specific to PTC (**Supplementary Figure 4**).

### Replication of the novel locus at 1p31.3

The lead SNP at 1p31.3, rs334729 lies in intron 3 of the *NFIA* gene (**Supplementary Figure 5**.**a**). This gene encodes a member of the NF1 (nuclear factor 1) family of transcription factors, and NF1 isoforms were shown to interact with thyroid transcription factor 1 (TTF-1) to regulate cell function in experimental studies (35). This locus was previously reported in a GWAS on thyroid stimulating hormone (TSH) levels (24,36) but not in GWAS on DTC. We conducted a replication analysis of rs334729 using data from two independent case-control studies conducted in Ohio (USA) (28) and Italy (25) totaling 2,270 cases and 2,125 controls and the association was nominally replicated (P=0.03) with the association in the same direction, but with significant heterogeneity between the three studies (P-heterogeneity=8.6×10^−3^) (**Supplementary Table 3**). Rs334729[C] is highly correlated with rs334725[C] (r^2^ =0.93) which has been found associated with decreased TSH levels in a GWAS conducted in an Islandic population (effect of rs334725 [C] = -0.13, P=1.0×10^−20^) (24). In that study, rs334725[C] was additionally analyzed in association with DTC in four studies which reported an increased risk (OR=1.30, P=6.6×10^−3^ for rs334725 [C]). We combined the results from these four studies with our findings in Europeans (OR=1.89, P=1.4×10^−7^ for rs334725[C]) in a meta-analysis and found an OR=1.52, P=6.1×10^−8^ (**Supplementary Table 4**) which was at the limit of the significance level.

### Replication of suggestive loci

All chromosomal regions with at least two SNPs associated with DTC at suggestive level P-value<10^−5^ in the GWAS conducted in Europeans (**Supplementary Table 3)** or in the pooled GWAS **(Supplementary Table 5**) were considered for replication. Due to the lack of replication sample, we did not attempt to replicate our findings in the Oceanian population. In addition to the known loci at 2q35, 8p12, 9q22.33, 14q13.3 and the novel locus 1p31.3, we identified 9 loci in Europeans and 11 loci in the pooled analysis corresponding to these criteria. The lead SNPs at each locus of interest were analyzed in the replication sample, and only variants at 5p15.33, 16q23.2 and 19p12 were associated with DTC at a nominal P-value <0.05 (**Supplementary Table 3** and **Supplementary Table 5**). For these SNPs, we report the results of the combined analysis of the three studies in European and the association in Oceanians in **Table 3**. Only the variants at 16q23.2 (rs16950982 and rs17767383) reached the genome-wide P-value threshold of 5×10^−8^ in the meta-analysis. Association with rs16950982[G] was in the same direction in both ethnic groups, while we observed opposite effects for the effect allele [A] of rs17767383 (**Table 3**). These two variants were in strong linkage disequilibrium (LD) in Europeans (r^2^ =0.88), but only in weak LD in Oceanians (r^2^ =0.37), and were located in an intergenic region close to the proto-oncogene *MAF* encoding a transcription factor (**Supplementary Figure 5**.**c**). Interestingly, rs16950982[G] was correlated with rs3813582[C] (r^2^ =0.88) which was previously associated with decreased TSH levels (β=-0.082, P=8.45×10^−18^) with a male-specific effect (36) also observed in our study on DTC (**Supplementary Figure 4**). Rs16950982[G] is also in LD with rs17767419[T] (r^2^ =0.88) which was associated with increased thyroid volume (β= 0.068, P=9.4×10-15) (37). According to GTEx Portal (https://www.gtexportal.org/home/), rs16950982[G] and rs17767383[A] are significantly associated with lower expression of *MAFTRR* in many tissues including thyroid tissue (respectively β=-0.55, P=2.8×10^−36^ and β=-0.60, P=2.7×10^−43^) and this gene is suspected to have a tumor-suppressor function (38).

**Table 3.**
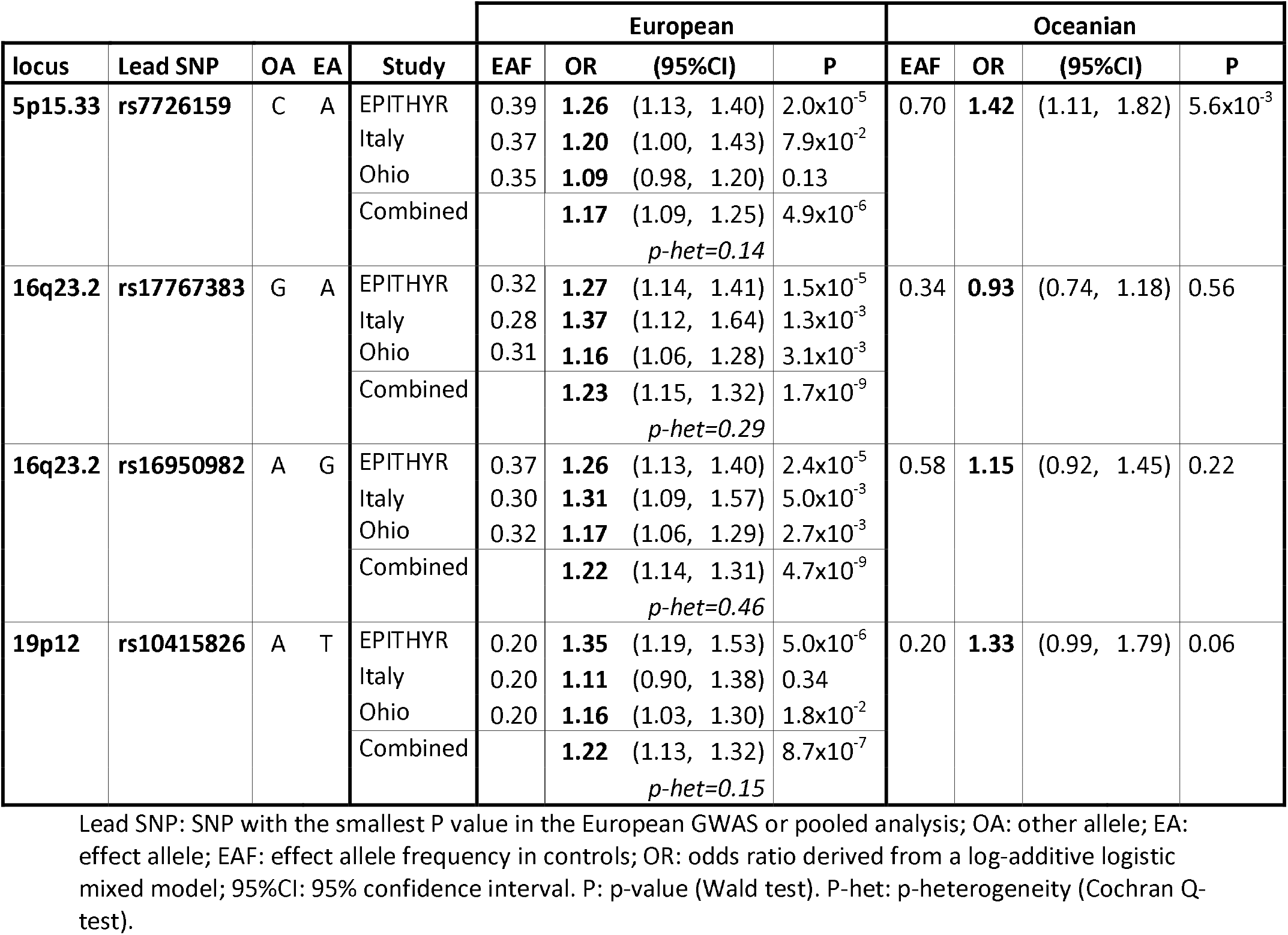
Replication analysis of suggestive loci.

At 5p15.33, we nominally replicated results for rs7726159 located in intron 3 of *TERT* (**Supplementary Figure 5**.**b**), but the combined association did not reach the GWAS significance threshold (P=4.9×10^−6^) (**Table 3, Supplementary Table 4**). We found that the associations in both ethnic groups were in the same direction but the allele frequency differed greatly, with a higher frequency of the risk allele [A] in the Oceanian ancestry population. In the meta-analysis of DTC GWAS in an European population (28), the best hit reported at 5p15.33 was for rs10069690[T] (OR=1.20, P=3.2×1^-7^), which was moderately correlated with our lead SNP rs7726159 (r^2^=0.46). Interestingly, another study conducted in a Chinese population (39) and analyzing 15 haplotype-tagging SNPs in the *TERT-CLPTM1* region reported an increased risk of PTC with rs2736100[G], which is highly correlated with rs7726159[A] in Asians (r^2^ =0.92 in 1000 genomes CHB population), but only moderately correlated in European populations (r^2^ =0.43 in 1000 genomes CEU population). Rs7726159[A] was also previously associated with increased telomere length in European (40) and Asian (41) populations, as well as with risk of multiple cancers including breast and ovarian cancers (40,42). This variant was also shown to enhance *TERT* transcription (41). At 19p12, we reported an association with rs10415826 (P=8.7×10^−7^) (**Table 3**), in intron 1 of *ZNF257* (**Supplementary Figure 5**.**d**). Little is known about the function of this gene encoding a transcription factor from the zinc finger family and to our knowledge, no association with other traits has been reported so far with this variant.

### Replication study of hits identified in a previously published meta-analysis of GWAS on DTC

The meta-analysis of DTC GWAS conducted by Gudmundsson et al. (28) identified five susceptibility loci at 1q42.2, 3q26.2, 5q22.1, 10q24.33 and 15q22.33, but the authors could not replicate these results in an independent sample. Here, we conducted a replication analysis of the lead SNPs at these loci and observed association with DTC in the same direction in Europeans for all SNPs; for SNPs at 1q42.2, 5q22.1 and 10q24 the P-values were <0.001. In Oceanians, associations with these SNPs were all in the opposite direction and none were significant (**Supplementary Table 6)**.

## DISCUSSION

EPITHYR constitutes a new consortium to study DTC risk factors; it gathers over 2,000 DTC cases and matched population controls of various geographical origin, with detailed epidemiological and genetic data. While lifestyle risk factors have been extensively analyzed in individual studies, this is the first analysis of genome-wide data in this consortium (10,20,50,31,43–49).

In summary, we confirmed the association with the known DTC susceptibility loci at 2q35, 8p12, 9q22.33 and 14q13.3 in the European ancestry population. We additionally replicated the association between DTC and a SNP at 5p15.33 (rs7726159) that correlated with rs2736100 which was reported in a previous Chinese study on PTC (39). Interestingly, the risk allele [A] of rs7726159 has also been associated with longer telomere length (40,41). This is in line with a Danish study in 95,568 individuals reporting that genetic determinants of long telomeres were associated with increased risk of cancer. In this study, it was hypothesized that long telomeres could lead to the development of cancers by promoting the survival of precancerous cells (42). Our results also suggest two novel DTC susceptibility loci at 1p31.3 (rs334729) and 16q23.2 (rs16950982), and the alleles of the SNPs conferring a higher risk of DTC have also been associated with decreased TSH levels in a previous GWAS (24,36). These results are concordant with previous epidemiological studies that measured thyroid hormones level in blood samples of DTC cases and their matched controls (51). Interestingly, the association with rs16950982 at 16q23.2 was stronger in men, which is also consistent with the results of the GWAS on TSH for this locus (36). Our study is the first GWAS conducted in an Oceanian population from French Polynesia and New Caledonia, two territories where the incidence of DTC is among the highest. Except at 1p31.3, all associations we highlighted in Europeans were in the same direction in the Oceanian ancestry population. However, our study was limited by the sample size and by the lack of replication sample in this population. Interestingly, we observed that the frequency of the risk alleles at 2q35, 5p15.33 and 16q23.2 were significantly higher in Oceanians than in Europeans (supplementary Table 7). This may explain part of the difference in DTC incidence observed between these two populations but additional GWAS and epidemiological studies in Oceanian populations are needed to fully understand the highest incidence observed in these populations.

## METHODS

### Discovery sample

The EPITHYR consortium included data from seven population-based case-control studies. Of these studies, three were conducted in Metropolitan France (CATHY, YOUNG-Thyr and E3N studies), two in South Pacific Islands (French Polynesia and New Caledonia), one in Cuba and one in Gomel region of Belarus contaminated after the Chernobyl fallout. The study designs are briefly outlined below and were described in detail previously (10,12,19,31,46,52). The number of participants in each study, as well as the number of participants for which a saliva or blood sample was available are shown in **Supplementary Table 1**.

All studies provided information on ethnicity, personal and familial history of thyroid disease, weight, height, dietary habits, alcohol intake, tobacco smoking, residential and occupational histories, and for women, menstrual and reproductive factors and exogenous hormone use. The EPITHYR sample constitutes the largest study on DTC including detailed epidemiological data and genetic data of the participants. All participants provided informed consent and each study was approved by their governing ethics committee.

### CATHY study

The CATHY study is a population-based case-control study performed in Marne, Ardennes, and Calvados, three French administrative areas (“départements”) covered by a cancer registry (19). Cases were patients aged 25 years and older who were diagnosed with DTC between 2002 and 2007 and residing in these areas. Controls matched to the cases by sex, age (5-year age groups), and study area were recruited by selecting telephone numbers at random. To prevent possible selection bias arising from differential participation rates across categories of socioeconomic status (SES) among controls, quotas by SES were also applied to reflect the distribution by SES categories in the general population. From 621 cases and 706 controls recruited for the study, saliva DNA samples (Oragene®) were obtained for 516 cases and 569 controls.

### New Caledonia (NC) study

The NC study is a country-wide, population-based, case-control study (10). The cases included patients with DTC diagnosed between 1993 and 1999 who had been living in NC for at least 5 years at the time of diagnosis. The cases were identified from the two pathology laboratories in NC and from active searches in the medical records of the main hospitals. Age- and sex-matched controls were randomly selected from recently updated electoral rolls. A total of 332 cases and 412 controls were included. Saliva DNA samples (Oragene®) were available for 224 cases and 261 controls.

### Young-Thyr Study

The Young-Thyr study population consists of 805 DTC cases and 876 cancer-free controls (46). The case-control study was conducted with patients younger than 35 years of age and born after January 1st, 1971, thereby excluding those who were older than 15 years of age at the time of the Chernobyl nuclear power plant accident. These cases were newly diagnosed with DTC between January 1st, 2002, and December 31st, 2006, and have their main residence at the time of diagnosis in a region of eastern France (Alsace, Champagne-Ardennes, Corse, Franche-Comté, Lorraine, Rhône-Alpes, and Provence-Alpes-Côte d’Azur). Controls were selected from the general population and individually-matched to a single case of the same sex and year of birth (within one year) and region of residence during the year when the case was diagnosed with cancer. They were randomly selected in each region from the landline telephone subscriber directory. DNA samples from saliva (Oragene®) were available for 715 cases and 692 controls.

### Cuban Study

The Cuban study population consists of 199 cases and 193 controls (31). All patients aged between 17 and 60 years, who were living in Havana and its surrounding municipality of Jaruco (30km from Havana), and were treated for DTC between 2000 and 2011 at INOR (National Institute of Oncology and Radiobiology) and at the Institute of Endocrinology were eligible for the study. Potential cases were selected from the National Cancer Registry databases and were cross-referenced with the INOR Pathology Register. Controls were selected from the general population living in Havana city and its surrounding municipality of Jaruco using consultation files from primary care units (family doctors). They were frequency-matched with cases by age at cancer diagnosis of the cases (±5 years) and gender. The study was approved by the Clinical Research Ethics Committee of the National Institute of Oncology and Radiobiology (INOR), Havana, Cuba.

### French Polynesia Study

The French Polynesia study population consists of 144 cases and 231 controls (53). All patients diagnosed with DTC, between 1983 and 2003, before the age of 56 years of age and born and living in French Polynesia were eligible for the study. Prevalent cases were identified from the cancer registry of French Polynesia, medical insurance files, and the four endocrinologists in Tahiti. Controls were individually matched on date of birth (+/- 6 months) and sex and were selected randomly from the French Polynesia registry of births, which records all inhabitants born in French Polynesia. The study was approved by the French Polynesian Ethics Committee.

### Chernobyl study

The study was conducted on a subgroup of subjects from the population-based case–control study carried out in the most contaminated areas of Belarus to evaluate the risk of thyroid cancer after exposure to radioactive iodine in childhood (52). Subjects included in the genetic study consisted of 83 PTC cases and 324 matched controls who consented to give a blood sample. Cases and controls included in the study were not related. All subjects were from the Gomel region in Belarus, and were younger than 15 years at the time of the Chernobyl accident. The control subjects were matched to the PTC cases by age (within 1 year for those who were 18 months or older at the time of the accident; within 6 months for those who aged 12–18 months and within 1 month for those who were younger than 12 months), sex and type of settlement. The cases were diagnosed within 6–12 years after the accident with histologically verified PTC confirmed by the International panel of pathologists. Two-thirds of the cases developed PTC before the age of 15 and the remaining cases before the age of 25. For more than 60% of cases, the latency (time between exposure and diagnosis) was less than 10 years.

The study was carried out with the approval of the IARC ethics committee and of the Belarus Coordinating Council for Studies of the Medical Consequences of the Chernobyl Accident.

Peripheral blood samples (10 ml) were collected in the presence of anticoagulant (sodium citrate) and processed for genomic DNA extraction using a standard inorganic method.

### E3N study

The E3N is a prospective cohort that included 98,995 women in 1990 who were living in France, born between 1925 and 1950 and covered by the MGEN, a National Health Insurance Plan for teachers and coworkers (12). Participants, who gave written informed consent for external health follow-up through the health insurer, completed biennial self-administered questionnaires, requesting information on lifestyle characteristics and health conditions. Blood samples were collected between 1994 and 1999 from 25,000 E3N participants and collection of saliva samples was conducted between 2010 and 2011 and were obtained from 47,000 participants. Participants were asked to report any diagnosis of cancer on each Q1 to Q8 questionnaire, and cases were then further explored by requesting information from the subjects’ physicians. Pathology reports were obtained for all incident thyroid cancer cases. For the present study, we considered only first primary DTC cases (i.e., PTC or FTC). For this study, sufficient DNA material was available for 568 participants: 372 DNA samples were extracted from saliva and 196 from buffy-coat.

### Replication sample

The Italian study included 701 cases consecutively recruited at the Department of Endocrinology of the University Hospital of Pisa, Italy. Controls (n=499) were healthy subjects recruited among blood donors and workers of the University Hospital of Pisa during their routine visits in the context of a program of surveillance performed by the Occupational Medicine unit. The eligibility criteria were the same for cases and controls, and it included a minimal age of 18 years. Individuals affected by any malignancy, chronic inflammatory disease, or related diseases in the past were excluded from the control group. A further exclusion criterion for controls was the presence of thyroid nodules or other benign thyroid diseases, when known. Both cases and controls should not have familial relationship with each other. All participants were of Caucasian origins. Other details of the study were published with the results of the GWAS (19). According to the Helsinki declaration, both healthy and affected volunteers gave their written informed consent to participate in the study and the study protocol was cleared by the local Ethical Committee.

The Ohio (USA) study is a case-control study that included 1,580 of self-reported European descent participants, including histologically confirmed PTC or FTC patients and 1,628 controls from the central Ohio area. Cases had a mean age of diagnosis of 43 years (74% are females) and controls had a mean age of 45 years at recruitment (74% are females). Genomic DNA was extracted from blood using standard procedures. The study was approved by the Institutional Review Board of the Ohio State and all participants provided written informed consent.

### Genotype data

#### OncoArray SNPs selection

All EPITHYR subjects were genotyped at the Centre National de Recherche en Génomique Humaine (CNRGH/CEA, Evry, France) using the Infinium OncoArray-500K BeadChip (Illumina), on an Illumina automated high-throughput platform. This array, which was previously described in details (54) contains 499,170 SNPs with a genome-wide backbone of about 250,000 tag SNPs designed such that the large majority of common variants could be accurately imputed. Additional SNPs include dense coverage across known genetic variants associated with breast, colorectal, lung, ovarian, and prostate cancers, SNPs covering ancestry, putative rare susceptibility variants from sequencing studies and other variants of potential biological relevance. As the OncoArray was not originally designed to include thyroid cancer susceptibility variants, we incorporated 13,759 additional custom markers based on prior evidence of association, candidate submissions, fine-mapping of known susceptibility regions and in genes involved in relevant biological pathways.

#### Quality control of genotyping data

Details of the genotyping calling for OncoArray are described in more details elsewhere (54). Cluster files generated using 56,284 samples from the GAME-ON consortium (available at http://consortia.ccge.medschl.cam.ac.uk/oncoarray/onco_v2c.zip) were applied to our dataset.

A total of 4,553 individuals were genotyped. We excluded samples with a call rate <95% (n=17), with a sex discordant call (n=12), and duplicates within and across studies (n=20). Additionally, we excluded pairs of individuals which were identified as duplicates using the genetic data but with different epidemiological data (n=10).

Ancestry was computed using the Fastpop program (55) which derived principal component analysis (PCA) scores from 2,318 informative markers and a subset of ∼47,000 individuals from the GAME-ON consortium. These scores were applied to our dataset. Individuals of European ancestry and Asian ancestry were defined as individuals an estimated proportion of European ancestry >0.8 and an estimated proportion of Asian ancestry >0.4, respectively with reference to the HapMap populations, based on the first two principal components (**Supplementary Figure 6**). Of the remaining 4,494 individuals, 3,538 were of European ancestry and 649 of Asian Ancestry. Additional individuals with extreme heterozygosity (4.89 standard deviation from the mean of the ethnicity) (n=11) were excluded and the final dataset comprised 3,527 individuals of European ancestry (1,554 cases and 1,973 controls) and 649 individuals of Asian ancestry (301 cases and 348 controls).

Of the 512,929 SNPs that were successfully manufactured, we received 493,266 SNPs with a call rate >90%. We excluded SNPs with a call rate <95% by study (n=8,327), duplicate SNPs (n=814), monomorphic SNPs (n=13,832), SNPs for which the cluster plots were judged to be not ideal (n=4,083). Of the remaining 466,210 SNPs, we additionally excluded SNPs not in HWE in each ethnic group (p<10^−7^ in controls, P<10^−12^ in cases) (n=563 in the study of European ancestry individuals and n=417 in the study of Asian ancestry individuals), monomorphic SNPs (n=5,210 in European ancestry individuals and n=37,513 in Asian ancestry individuals) as well as non-monomorphic SNPs with MAF<1% (n=58,912 in European ancestry individuals and n=75,177 Asian ancestry individuals). We retained for the analysis 401,525 SNPs among Europeans and 353,103 SNPs among Asians (**Supplementary Figure 7**).

#### Imputation

We performed the imputation of missing SNPs in each population (European and Asian) independently using the October 2014 (version 3) release of the 1000 Genomes Project dataset as the reference panel. We excluded from imputation (but not from association analysis) SNPs with a call<98% in each ethnic group, SNPs that could not be linked to the 1000 Genomes project reference as well as ambiguous SNPs (AT/GC).

For the European population, we additionally excluded SNPs for which the difference statistic (|p_1_-p_2_|-0.01)^2^/((p_1_ +p_2_)(2-p_1_ -p_2_)) was > 0.008, where p1 and p2 are respectively the frequencies in our data set and in the 1000 Genomes Project. Therefore, imputation in Europeans was based on 362,025 genotyped SNPs and a two-stage procedure was performed, using SHAPEIT2 (using the default parameters (“—burn 10 –prune 10 –main 50”) to derive phased genotypes and IMPUTEv2 to perform imputation using 5 Mb non-overlapping intervals. The number of reference haplotypes to use as templates when imputing missing genotypes was increased to 800 (“-k_hap 800”), and the buffer region was increased to 500kb (“-buffer 500”). We excluded imputed SNPs with an imputation r^2^<0.3 and SNPs with a MAF <1% in our population leaving a total of 9,673,063 SNPs.

For the Asian population, we imputed the non-genotyped data with IMPUTEv2 but without pre-phasing in SHAPEIT, which is computationally more demanding but improves imputation accuracy (56). Because our “Asian” population was closer to Oceanian populations (**Supplementary Figure 8**), we added to the 1000 Genomes project reference panel, genomes from 26 Oceanian individuals from the Simons Diversity Project that were available publicly (https://sharehost.hms.harvard.edu/genetics/reich_lab/sgdp/phased_data/). Imputation was based on 314,954 SNPs. We excluded SNPs with an imputation r^2^<0.3 and SNPs with a MAF <1% in our population leaving a total of 7,179,638 SNPs.

### Statistical analysis

Odds ratios (OR) were calculated using logistic mixed models with the genetic relationship matrix as random effect to account for population structure and relatedness (57). All analyses were adjusted for age, sex, and study. We conducted separate analysis in Europeans and Oceanians, as well as a pooled analysis combining both populations. We also performed stratified analyses by sex, age group (<50, ≥50 years-old), histology (papillary/follicular cancer), and size of carcinoma for papillary cancers (<10mm/≥10 mm). We used the GASTON R package (https://cran.r-project.org/web/packages/gaston/index.html) to perform the logistic mixed models. For genome-wide analyses, P-values for each genetic variant were derived from a score test using a logistic mixed model as this test is less computationally demanding than a Wald test but asymptotically equivalent. A Wald test was then performed for a subset of variants of interest in order to get the effect size estimate. The traditional Bonferroni threshold for GWAS at P<5×10^−8^ was used to assess significance.

In the replication phase, we performed a fixed effect meta-analysis with inverse-variance weighting as implemented in METAL software (58). Heterogeneity of ORs across the studies and across the stratification groups was assessed using the Cochran’s Q-test.

## Data Availability

The data that support the findings of this study are available on request from the corresponding author. The data are not publicly available due to privacy or ethical restrictions.

## Acknowledgements

The EPITHYR GWAS received financial support from INCA (grant number 9533) and ARC (grant number PGA120150202302). The participating studies were funded by Ligue Nationale Contre le Cancer, Agence Nationale pour la Recherche (ANR), the Direction Générale de la Sante, the Agence Française de Sécurité Sanitaire de l’alimentation, de l’environnement et du travail (ANSES), CHILDTHYR EEC program, and the Fondation de France. JG was the recipient of a PhD fellowship from Région Ile-de-France. We thank Dr. Yannick Rougier and Dr. Dominique Baron-Dubourdieu for providing pathological reports for the NC study, as well as Dr. Sylvie Laumond, Dr. Jean-Paul Grangeon (Direction des affaires sanitaires et sociales de Nouvelle-Calédonie) and the country’s provincial health authorities (DPASS Sud, DPASS Nord, DPASS Iles Loyauté) for support during data collection in the NC study. We thank Milagros Velasco, Mae Chappe, and Idalmis Infante (Institute of Oncology and Radiobiology, La Havana, Cuba) and Silvia Turcios (National Institute of Endocrinology, La Havana, Cuba), for helping in the collection of the data in the Cuban Study. We thank John Paoaafaite and Joseph Teuri, who contacted and interviewed cases and controls for the study. Finally, we also thank P. Morales, J. Iltis, P. Giraud, P. Didiergeorge, M. Brisard, G. Soubiran, B. Caillou, P. Dupire, J. Ienfa, G. de Clermont, N. Cerf, B. Oddo, M. Bambridge, C. Baron, A. Mouchard-Rachet, O. Simonet, D. Lamarque, J. Vabret, J. Delacre, M.P. Darquier, and J. Leninger, for their help in the collection of the cases or in the organization of study in French Polynesia. We would like to thank the Association Centre de Regroupement Informatique et Statistique en Anatomie Pathologique en Provence-Alpes-Côte d’Azur (CRISAP PACA), as well as Dr. Arlette Danzon, Dr. Geneviève Sasolas, Dr. Marc Christophe Sattonnet, Dr. Marc Colonna, Dr Brigitte Lacour, Dr Michel Velten, Dr. Enora Clero, Dr. Stephane Maillard, Dr. Laurent Bailly, Dr. Eugènia Mariné Barjoan, Dr Jean- Luc Lassalle, Dr Z Hafdi- Nejjari, Dr P Delafosse, Dr Elisabeth Adjadj, Kami-Marie Moreau, Cyrielle Orenes, Laurianne Sarrazin, Stéphanie Bonnay, Frédérique Chatelain, Maryse Barouh, Evelyne Rapp, Julie Festraëts, Julie Valbousquet, YusufAtilgan, Jean Chappellet, Lallia Bedhouche, Florent Dayet and Ziyan Fami, for their help in the collection of cases, the organization and the management of the Young-Thyr study. We acknowledge Stefano Landi for the Italian GWAS data and Subhayan Chattopadhyay and Yasmeen Niazi (Division of Molecular Genetic Epidemiology, German Cancer Research Center - DKFZ) for technical assistance in these data analysis. We are grateful to Dr. Hervé Perdry for his help in using the GASTON package.

## Author contributions

Project coordination: TT, FL, FDV, PG. Selection of add-ons on the Oncoarray chip: TT, FL, CX, CR, FDV. Quality controls and data management was done by JG, MK, PE under the supervision of TT. Genotyping was supervised and carried out by DB, AB and JFD. Imputation strategy was proposed by AFH, ALL, TT. Bioinformatics analysis were performed by PE, MK, OK, EL. Statistical analysis was carried out by PE, TT. Subject recruitment, biological material collection and handling was organized and carried out by FDV, PG, CS, AVG, CM, PLP, MCB, AK, RMO, JJLA, CX, YR, FBC, FR, TT. Summary statistics of replication samples were provided by ADC, DC, SL, HH, AF, HT, FG, RE. TT, FL and PE drafted the manuscript. All authors reviewed the manuscript and approved the final version of the paper.

## Disclaimer

Where authors are identified as personnel of the International Agency for Research on Cancer / World Health Organization, the authors alone are responsible for the views expressed in this article and they do not necessarily represent the decisions, policy or views of the International Agency for Research on Cancer / World Health Organization

## References

1. Bray F, Ferlay J, Soerjomataram I, Siegel R, Torre L, Jemal A. Global cancer statistics 2018: GLOBOCAN estimates of incidence and mortality worldwide for 36 cancers in 185 countries. CA Cancer J Clin. France: In. 2018;68(6):394–424.

2. Magreni A, Bann D V., Schubart JR, Goldenberg D. The effects of race and ethnicity on thyroid cancer incidence. JAMA Otolaryngol - Head Neck Surg. 2015;141(4):319–23.

3. Blot W, Le Marchand L, Boice JJ, Henderson BE. Thyroid cancer in the Pacific. JNCI J Natl Cancer Inst. 1997;89(1):90–1.

4. Truong T, Rougier Y, Dubourdieu D, Guihenneuc-Jouyaux C, Orsi L, Hémon D, et al. Time trends and geographic variations for thyroid cancer in New Caledonia, a very high incidence area (1985–1999). Eur J Cancer Prev. 2007 Feb;16(1):62–70.

5. Meredith I, Sarfati D, Atkinson J, Blakely T. Thyroid cancer in Pacific women in New Zealand. N Z Med J. 2014 Jun 6;127(1395):52–62.

6. Wiltshire JJ, Drake TM, Uttley L, Balasubramanian SP. Systematic Review of Trends in the Incidence Rates of Thyroid Cancer. Thyroid. 2016;26(11):1541–52.

7. Kitahara CM, Sosa JA. The changing incidence of thyroid cancer. Nat Rev Endocrinol. 2016 Nov;12(11):646–53.

8. Iglesias ML, Schmidt A, Ghuzlan A Al, Lacroix L, Vathaire F de, Chevillard S, et al. Radiation exposure and thyroid cancer: a review. Arch Endocrinol Metab. 2017 Mar;61(2):180–7.

9. Wang H. Obesity and Risk of Thyroid Cancer: Evidence from a Meta-Analysis of 21 Observational Studies. Med Sci Monit. 2015;21:283–91.

10. Guignard R, Truong T, Rougier Y, Baron-Dubourdieu D, Guénel P. Alcohol drinking, tobacco smoking, and anthropometric characteristics as risk factors for thyroid cancer: A countrywide case-control study in New Caledonia. Am J Epidemiol. 2007;166(10):1140–9.

11. Brindel P, Doyon F, Rachédi F, Boissin J-L, Sebbag J, Shan L, et al. Anthropometric factors in differentiated thyroid cancer in French Polynesia: a case-control study. Cancer causes Control. 2009 Jul;20(5):581–90.

12. Clavel-Chapelon F, Guillas G, Tondeur L, Kernaleguen C, Boutron-Ruault MC. Risk of differentiated thyroid cancer in relation to adult weight, height and body shape over life: The French E3N cohort. Int J Cancer. 2010;126(12):2984–90.

13. Lee JH, Hwang Y, Song RY, Yi JW, Yu HW, Kim SJ, et al. Relationship between iodine levels and papillary thyroid carcinoma: A systematic review and meta-analysis. Head Neck. 2017;39(8):1711–8.

14. Zimmermann MB, Galetti V. Iodine intake as a risk factor for thyroid cancer: A comprehensive review of animal and human studies. Thyroid Res. 2015;8(1):1–21.

15. Caini S, Gibelli B, Palli D, Saieva C, Ruscica M, Gandini S. Menstrual and reproductive history and use of exogenous sex hormones and risk of thyroid cancer among women: a meta-analysis of prospective studies. Cancer Causes Control. 2015;26(4):511–8.

16. Cho YA, Kim J. Thyroid cancer risk and smoking status: a meta-analysis. Cancer Causes Control. 2014 Sep;25(9):1187–95.

17. Hong S-H, Myung S-K, Kim H. Alcohol Intake and Risk of Thyroid Cancer: A Meta-analysis of Observational Studies. Cancer Res Treat. 2016 Jul;

18. Wu L, Zhu J. Linear reduction in thyroid cancer risk by oral contraceptive use: A dose-response meta-analysis of prospective cohort studies. Hum Reprod. 2015;30(9):2234–40.

19. Cordina-Duverger E, Leux C, Neri M, Tcheandjieu C, Guizard AV, Schvartz C, et al. Hormonal and reproductive risk factors of papillary thyroid cancer: A population-based case-control study in France. Cancer Epidemiol. 2017;48:78–84.

20. Xhaard C, Rubino C, Cléro E, Maillard S, Ren Y, Borson-Chazot F, et al. Menstrual and reproductive factors in the risk of differentiated thyroid carcinoma in young women in France: a population-based case-control study. Am J Epidemiol. 2014 Nov 15;180(10):1007–17.

21. Hemminki K, Li X. Familial risk of cancer by site and histopathology. Int J Cancer. 2003;103(April 2002):105–9.

22. Gudmundsson J, Sulem P, Gudbjartsson DF, Jonasson JG, Sigurdsson A, Bergthorsson JT, et al. Common variants on 9q22.33 and 14q13.3 predispose to thyroid cancer in European populations. Nat Genet. 2009 Apr;41(4):460–4.

23. Takahashi M, Saenko V a, Rogounovitch TI, Kawaguchi T, Drozd VM, Takigawa-Imamura H, et al. The FOXE1 locus is a major genetic determinant for radiation-related thyroid carcinoma in Chernobyl. Hum Mol Genet. 2010 Jun 15;19(12):2516–23.

24. Gudmundsson J, Sulem P, Gudbjartsson DF, Jonasson JG, Masson G, He H, et al. Discovery of common variants associated with low TSH levels and thyroid cancer risk. Nat Genet. 2012 Mar;44(3):319–22.

25. Köhler A, Chen B, Gemignani F, Elisei R, Romei C, Figlioli G, et al. Genome-wide association study on differentiated thyroid cancer. J Clin Endocrinol Metab. 2013 Oct;98(10):E1674–81.

26. Figlioli G, Chen B, Elisei R, Romei C, Campo C, Cipollini M, et al. Novel genetic variants in differentiated thyroid cancer and assessment of the cumulative risk. Sci Rep. 2015;5:8922.

27. Mancikova V, Cruz R, Inglada-Pérez L, Fernández-Rozadilla C, Landa I, Cameselle-Teijeiro J, et al. Thyroid cancer GWAS identifies 10q26.12 and 6q14.1 as novel susceptibility loci and reveals genetic heterogeneity among populations. Int J Cancer. 2015;137(8):1870–8.

28. Gudmundsson J, Thorleifsson G, Sigurdsson JK, Stefansdottir L, Jonasson JG, Gudjonsson SA, et al. A genome-wide association study yields five novel thyroid cancer risk loci. Nat Commun. 2017;8:14517.

29. Son H-Y, Hwangbo Y, Yoo S-K, Im S-W, Yang SD, Kwak S-J, et al. Genome-wide association and expression quantitative trait loci studies identify multiple susceptibility loci for thyroid cancer. Nat Commun. 2017;8(May):15966.

30. Tcheandjieu C, Lesueur F, Sanchez M, Baron-Dubourdieu D, Guizard AV, Mulot C, et al. Fine-mapping of two differentiated thyroid carcinoma susceptibility loci at 9q22.33 and 14q13.3 detects novel candidate functional SNPs in Europeans from metropolitan France and Melanesians from New Caledonia. Int J Cancer. 2016;

31. Lence-Anta JJ, Xhaard C, Ortiz RM, Kassim H, Pereda CM, Turcios S, et al. Environmental, lifestyle, and anthropometric risk factors for differentiated thyroid cancer in Cuba: A case-control study. Eur Thyroid J. 2014;3:189–96.

32. Wang Y-L, Feng S-H, Guo S-C, Wei W-J, Li D-S, Wang Y, et al. Confirmation of papillary thyroid cancer susceptibility loci identified by genome-wide association studies of chromosomes 14q13, 9q22, 2q35 and 8p12 in a Chinese population. J Med Genet. 2013 Oct;50(10):689–95.

33. Maillard S, Damiola F, Clero E, Pertesi M, Robinot N, Rachédi F, et al. Common variants at 9q22.33, 14q13.3, and ATM loci, and risk of differentiated thyroid cancer in the French Polynesian population. PLoS One. 2015;10(4):1–14.

34. Pereda C, Lesueur F, Pertesi M, Robinot N, Lence-Anta JJ, Turcios S, et al. Common variants at the 9q22.33, 14q13.3 and ATM loci, and risk of differentiated thyroid cancer in the Cuban population. BMC Genet. 2015;16(22).

35. Bachurski CJ, Yang GH, Currier TA, Gronostajski RM, Hong D. Nuclear Factor I/Thyroid Transcription Factor 1 Interactions Modulate Surfactant Protein C Transcription. Mol Cell Biol. 2003;23(24):9014–24.

36. Porcu E, Medici M, Pistis G, Volpato CB, Wilson SG, Cappola AR, et al. A Meta-Analysis of Thyroid-Related Traits Reveals Novel Loci and Gender-Specific Differences in the Regulation of Thyroid Function. PLoS Genet. 2013;9(2).

37. Teumer A, Rawal R, Homuth G, Ernst F, Heier M, Evert M, et al. Genome-wide association study identifies four genetic loci associated with thyroid volume and goiter risk. Am J Hum Genet. 2011;88(5):664–73.

38. Cheng J, Demeulemeester J, Wedge DC, Vollan HKM, Pitt JJ, Russnes HG, et al. Pan-cancer analysis of homozygous deletions in primary tumours uncovers rare tumour suppressors. Nat Commun. 2017;8(1):1–14.

39. Ge M, Shi M, An C, Yang W, Nie X, Zhang J, et al. Functional evaluation of TERT-CLPTM1L genetic variants associated with susceptibility of papillary thyroid carcinoma. Sci Rep. 2016;6(May):1–7.

40. Pooley KA, Bojesen SE, Weischer M, Nielsen SF, Thompson D, Amin Al Olama A, et al. A genome-wide association scan (GWAS) for mean telomere length within the COGS project: Identified loci show little association with hormone-related cancer risk. Hum Mol Genet. 2013;22(24):5056–64.

41. Li Y, Xiang C, Shen N, Deng L, Luo X, Yuan P, et al. Functional polymorphisms on chromosome 5p15.33 disturb telomere biology and confer the risk of non-small cell lung cancer in Chinese population. Mol Carcinog. 2019;58(6):913–21.

42. Rode L, Nordestgaard BG, Bojesen SE. Long telomeres and cancer risk among 95 568 individuals from the general population. Int J Epidemiol. 2016;45(5):1634–43.

43. Leux C, Truong T, Petit C, Baron-Dubourdieu D, Guénel P. Family history of malignant and benign thyroid diseases and risk of thyroid cancer: a population-based case–control study in New Caledonia. Cancer Causes Control. 2012;23:745–55.

44. Truong T, Orsi L, Dubourdieu D, Rougier Y, Hémon D, Guénel P. Role of goiter and of menstrual and reproductive factors in thyroid cancer: a population-based case-control study in New Caledonia (South Pacific), a very high incidence area. Am J Epidemiol. 2005 Jun;161(11):1056–65.

45. Michailidou K, Lindström S, Dennis J, Beesley J, Hui S, Kar S, et al. Association analysis identifies 65 new breast cancer risk loci. Nature. 2017;551(7678):92–4.

46. Xhaard C, De Vathaire F, Cléro E, Maillard S, Ren Y, Borson-Chazot F, et al. Anthropometric risk factors for differentiated thyroid cancer in young men and women from Eastern France: A casecontrol study. Am J Epidemiol. 2015;182(3):202–14.

47. Xhaard C, Ren Y, Clero E, Maillard S, Brindel P, Rachedi F, et al. Differentiated thyroid carcinoma risk factors in French Polynesia. Asian Pac J Cancer Prev. 2014 Jan;15(6):2675–80.

48. Truong T, Baron-Dubourdieu D, Rougier Y, Guénel P. Role of dietary iodine and cruciferous vegetables in thyroid cancer: A countrywide case-control study in New Caledonia. Cancer Causes Control. 2010;21:1183–92.

49. Cléro É, Doyon F, Chungue V, Rachédi F, Boissin J-L, Sebbag J, et al. Dietary iodine and thyroid cancer risk in French Polynesia: a case-control study. Thyroid. 2012 Apr;22(4):422–9.

50. Cardis E, Kesminiene A, Ivanov V, Malakhova I, Shibata Y, Khrouch V, et al. Risk of thyroid cancer after exposure to 131I in childhood. J Natl Cancer Inst. 2005 May 18;97(10):724–32.

51. Rinaldi S, Plummer M, Biessy C, Tsilidis KK, Ostergaard JN, Overvad K, et al. Thyroid-stimulating hormone, thyroglobulin, and thyroid hormones and risk of differentiated thyroid carcinoma: The EPIC study. J Natl Cancer Inst. 2014;106(6).

52. Damiola F, Byrnes G, Moissonnier M, Pertesi M, Deltour I, Fillon A, et al. Contribution of ATM and FOXE1 (TTF2) to risk of papillary thyroid carcinoma in Belarusian children exposed to radiation. Int J Cancer. 2014 Apr 1;134(7):1659–68.

53. Brindel P, Doyon F, Rachédi F, Boissin JL, Sebbag J, Shan L, et al. Anthropometric factors in differentiated thyroid cancer in French Polynesia: A case-control study. Cancer Causes Control. 2009;20:581–90.

54. Amos CI, Dennis J, Wang Z, Byun J, Schumacher FR, Gayther SA, et al. The OncoArray Consortium: a Network for Understanding the Genetic Architecture of Common Cancers. Cancer Epidemiol Biomarkers Prev. 2017;26(1):126–35.

55. Li Y, Byun J, Cai G, Xiao X, Han Y, Cornelis O, et al. FastPop: A rapid principal component derived method to infer intercontinental ancestry using genetic data. BMC Bioinformatics. 2016;17:122.

56. Roshyara NR, Horn K, Kirsten H, Ahnert P, Scholz M, An P, et al. Comparing performance of modern genotype imputation methods in different ethnicities. Sci Rep. 2016;6:34386.

57. Chen H, Wang C, Conomos MP, Stilp AM, Li Z, Sofer T, et al. Control for Population Structure and Relatedness for Binary Traits in Genetic Association Studies via Logistic Mixed Models. Am J Hum Genet. 2016;98(4):653–66.

58. Willer CJ, Li Y, Abecasis GR. METAL: Fast and efficient meta-analysis of genomewide association scans. Bioinformatics. 2010;26(17):2190–1.

